# US Mortality Due To Congenital Heart Disease Across the Lifespan from 1999-2017 Exposes Persistent Racial/Ethnic Disparities

**DOI:** 10.1101/2020.03.15.20036525

**Authors:** Keila N. Lopez, Shaine A. Morris, Kristen Sexson Tejtel, Andre Espaillat, Jason L. Salemi

## Abstract

**Background:** Congenital heart disease (CHD) accounts for approximately 40% percent of deaths in United States (US) children with birth defects. Previous US data from 1999-2006 demonstrated an overall decrease in CHD mortality. The objective of our study was to assess current trends in US mortality related to CHD from infancy to adulthood over the last 19 years and determine differences by sex and race/ethnicity.

**Methods:** We conducted an analysis of death certificates from 1999-2017 to calculate annual CHD mortality by age at death, race/ethnicity, and sex. Population estimates used as denominators in mortality rate calculation for infants were based on National Center for Health Statistics live birth data. Mortality rates in individuals ≥1 year of age utilized US Census Bureau bridged-race estimates as denominators for population estimates. We characterized temporal trends in all-cause mortality, mortality resulting directly due to and related to CHD by age, race/ethnicity, and sex using joinpoint regression.

**Results:** There were 47.7 million deaths with 1 in 814 deaths due to CHD (n=58,599). While all-cause mortality decreased 16.4% across all ages, mortality resulting from CHD declined 39.4% overall. The mean annual decrease in CHD mortality was 2.6%, with the largest decrease for those age >65years. The age-adjusted mortality rate decreased from 1.37 to 0.83 per 100,000. Males had higher mortality due to CHD than females throughout the study, although both sexes declined at a similar rate (∼40% overall), with a 3-4% annual decrease between 1999 and 2009, followed by a slower annual decrease of 1.4% through 2017. Mortality resulting from CHD significantly declined among all race/ethnicities studied, although disparities in mortality persisted for non-Hispanic Blacks versus non-Hispanic Whites (mean annual decrease 2.3% versus 2.6%, respectively; age-adjusted mortality rate 1.67 to 1.05 versus 1.35 to 0.80 per 100,000, respectively).

**Conclusions:** While overall US mortality due to CHD has decreased over the last 19 years, disparities in mortality persist for males compared to females and for non-Hispanic Blacks compared to non-Hispanic Whites. Determining factors that contribute to these disparities such as access to quality care, timely diagnosis, and maintenance of insurance will be important moving into the next decade.

## INTRODUCTION

Congenital heart disease is the leading cause of birth defect-related mortality, accounting for approximately 40% percent of deaths in children with birth defects in the United States and worldwide.^1–4^ Prior studies have demonstrated decreasing mortality over time in CHD in the United States, likely due to marked advances in medical and surgical care. ^5,6^ The most comprehensive assessment in the United States to date was a population-based study evaluating CHD mortality from 1999 through 2006.^5^ This study demonstrated that while decreasing mortality was noted across all race/ethnicities and both sexes, disparities persisted over time, with higher mortality described in Non-Hispanic (NH) Blacks compared to NH Whites and in males compared to females throughout the lifespan. Other studies have shown that the increased mortality for NH Black children has been predominantly in the lowest mortality CHD lesions, suggesting an influence of sociodemographic factors.^7,8^ While children of Hispanic ethnicity have not demonstrated the same disparities in CHD mortality in population-based studies, subgroup analysis of Hispanics with CHD in the United Stated demonstrated that infants of Mexican mothers had higher mortality than those of non-Hispanic White mothers.^9^

More recent population-based CHD mortality data are lacking. There have been continued advances in CHD care, including a dramatic increase in prenatal diagnosis of CHD, a push to prevent preterm and early term deliveries, national collaborative quality initiatives in perioperative care of CHD, and public reporting of many CHD outcomes. ^10–17^ There has also been increased attention to sex, racial/ethnic, and sociodemographic disparities in care of people affected by CHD across the age spectrum. ^18^ The objective of our study was to update prior data on temporal trends in mortality related to CHD in the United States by examining mortality over the last 19 years overall, by age and by CHD lesion, to determine if mortality is continuing to decline, and to evaluate if prior disparities noted across race/ethnicity and sex are decreasing.

## METHODS

### Mortality (numerator) data

We conducted a 19-year analysis of Multiple Cause of Death (MCOD) data files compiled and produced annually by the National Center for Health Statistics at the Centers for Disease Control and Prevention.^19^ As our study was based on analyses that used publicly available, de-identified MCOD and census data, it was deemed exempt by the Baylor College of Medicine Institutional Review Board. The MCOD files are based on death certificates filed by registries in all 50 states and the District of Columbia. Deaths among nonresident aliens, nationals living abroad, residents of US territories, and fetal deaths are excluded from MCOD files. Each record is based on a death certificate containing a single underlying cause of death (UCOD), up to 20 contributing causes of death, and demographic and geographic data on each decedent. The UCOD is assigned based on the World Health Organization’s definition of “*the disease or injury which initiated the train of morbid events leading directly to death, or the circumstances of the accident or violence which produced the fatal injury*.”^20^ As is the case for all deaths from 1999 and beyond, the UCOD and all listed contributing causes of death were captured using International Classification of Diseases, Tenth Revision (ICD-10) codes. For each decedent, we also ascertained sex (male, female), age in years (categorized as <1, 1 to 4, 5 to 17, 18 to 34, 35 to 49, 50 to 64, and ≥65), race/ethnicity, and associated CHD. Race and Hispanic origin are reported as separate variables on the death certificate; the final racial/ethnic category, defined to be consistent with categories used in bridged-race population estimates, ^21^ was first based on ethnicity (Hispanic or non-Hispanic [NH]) and the NH group was further subdivided by race (NH white, NH black, and NH other). Specific CHD lesion mortality was assessed overall in the lifespan, as well as in the infant period. Median age at death was also assessed for the study period.

In addition to deaths from any cause (all-cause mortality), we used the UCOD and contributing causes of death to define several CHD-specific mortality subgroups. Mortality resulting from (due to) CHD was defined as a death certificate in which the UCOD field contained an ICD-10 code in the Q20-Q26 range that is indicative of a CHD. CHD-related deaths were a broader category in which either the UCOD or any contributing cause of death had a CHD-related ICD-10 code.

### Population (denominator) data

Population estimates used as denominators in the calculation of mortality rates for decedents less than one year of age were based on live birth data downloaded from the National Center for Health Statistics.^22^ For decedents one year of age and older, population estimates used as denominators in the calculation of mortality rates were bridged-race estimates from the US Census Bureau.^21^ Population counts for 1999 were intercensal estimates based on the 1990 and 2000 decennial census; population counts for 2000 and 2010 were based on the decennial census conducted in their respective years; population counts for 2001-2009 were revised intercensal estimates based on the 2000 and 2010 decennial census; and population counts for 2011-2017 were postcensal estimates, combining the 2010 decennial census data with birth, death, migration, and net international immigration data.

### Statistical analyses

For each year of the study, and across strata of sex and race/ethnicity, we calculated age-specific death rates for 1) all-cause mortality, 2) mortality due to CHD, and 3) CHD-related mortality. Rates were calculated as the number of deaths per 1,000 live births for infants <1 year or per 100,000 population for ages ≥1 year. The population subgroups of sex and race/ethnicity whose mortality was being compared can differ in their age distributions, and age is strongly positively correlated with mortality. Therefore, to facilitate comparison across subgroups and from year to year, we also calculated age-adjusted mortality rates using the direct standardization method in which age-specific death rates were weighted according to the 2000 Standard Population.^23^

We assessed temporal trends in all-cause mortality, mortality resulting directly due to and related to CHD by age, race/ethnicity, sex, and specific CHD. Since the timeframe of the previous nationwide assessment of CHD mortality was 1999 to 2006,^5^ we presented death rates for 1999 and 2006, and added 2017, the most recent year of the current study. We then estimated and characterized temporal trends in mortality across the entire 19-year study period using joinpoint regression, which is particularly effective at identifying changes in the temporal trends of events over time.^24^ The first step in joinpoint regression is to fit annual mortality rate data to a straight line – one with zero joinpoints – that assumes a single trend best characterizes how rates change over the entire assessment period. Then, a joinpoint is added to the model at the most likely inflection point at which the trend changes, and a Monte Carlo permutation test is used to determine whether the added joinpoint improves model fit; if so, the joinpoint will be incorporated.^24^ The addition and assessment of additional joinpoints proceeds in an iterative fashion until a final model is selected with an optimal number of joinpoints. In the final model, each joinpoint corresponds to a statistically significant change in the mortality trend, and how mortality changes within each distinct time interval is characterized with an annual percent change (APC) and its associated 95% confidence interval (CI). Joinpoint regression also provides an estimate of the average APC (AAPC), which is a single value that describes trends in mortality over the entire study period, even when there are significant changes in the trend over time.^24^ To assess the extent to which race/ethnic disparities in all-cause and CHD mortality were changing during the study period, we calculated age-standardized mortality rate ratios (e.g., comparing NH blacks to NH whites) and infant mortality rate ratios for each year of the study.

To maintain consistency with previously published research on CHD-related mortality in the US, our base case analyses included all deaths associated with CHD. However, as a sensitivity analysis, in an attempt to assess CHD mortality independent of prematurity and extracardiac birth defects, we re-ran our analyses after excluding as a CHD-related death any case with either prematurity, an extracardiac defect, or genetic syndrome as a contributing cause of death (Supplemental Table 1).

**Table 1.** Temporal Trends in All-Cause Mortality and Mortality Resulting From CHD, by Age, Race/Ethnicity, and Sex, United States, 1999–2017

| Population subgroup | CHD listed as underlying cause of death |  |  |  | All-Cause |  |  |  |
| --- | --- | --- | --- | --- | --- | --- | --- | --- |
|  | 1999 | 2006 | 2017 | 99-17 AAPC<br>(95% CI) | 1999 | 2006 | 2017 | 99-17 AAPC<br>(95% CI) |
| **All ages** |  |  |  |  |  |  |  |  |
| Overall | 1.37 | 1.03 | 0.83 | -2.6 (-2.9, -2.2) | 875.20 | 791.26 | 732.07 | -1.1 (-1.2, -0.9) |
| Race/ethnicity |  |  |  |  |  |  |  |  |
| NH white | 1.35 | 1.01 | 0.80 | -2.6 (-3.0, -2.2) | 859.58 | 788.72 | 755.15 | -0.8 (-1.0, -0.7) |
| NH black | 1.67 | 1.41 | 1.05 | -2.3 (-2.8, -1.8) | 1149.35 | 1018.60 | 882.25 | -1.5 (-1.7, -1.3) |
| Hispanic | 1.20 | 0.86 | 0.81 | -2.2 (-2.8, -1.5) | 676.26 | 603.83 | 525.64 | -1.5 (-1.7, -1.3) |
| NH other | 0.85 | 0.73 | 0.55 | -2.9 (-3.4, -2.3) | 570.67 | 499.98 | 438.59 | -1.4 (-1.6, -1.2) |
| Sex |  |  |  |  |  |  |  |  |
| Male | 1.47 | 1.16 | 0.92 | -2.4 (-2.8, -2.0) | 1066.58 | 942.84 | 864.63 | -1.2 (-1.4, -1.1) |
| Female | 1.26 | 0.91 | 0.74 | -2.7 (-3.2, -2.3) | 733.61 | 671.75 | 619.86 | -1.0 (-1.2, -0.7) |
| **Age at death, <1 y** |  |  |  |  |  |  |  |  |
| Overall | 45.14 | 37.42 | 29.52 | -2.2 (-2.6, -1.8) | 704.86 | 667.58 | 577.92 | -1.0 (-1.4, -0.7) |
| Race/ethnicity |  |  |  |  |  |  |  |  |
| NH white | 44.18 | 35.12 | 26.89 | -2.6 (-2.9, -2.2) | 577.44 | 558.89 | 469.52 | -1.0 (-1.7, -0.3) |
| NH black | 56.70 | 53.77 | 40.26 | -1.8 (-2.3, -1.3) | 1458.00 | 1377.77 | 1199.01 | -1.0 (-2.2, 0.2) |
| Hispanic | 45.08 | 36.62 | 33.66 | -1.8 (-2.6, -1.1) | 574.89 | 548.40 | 530.69 | -0.5 (-0.7, -0.4) |
| NH other | 28.73 | 25.52 | 17.77 | -3.2 (-4.2, -2.3) | 463.49 | 430.20 | 334.07 | -1.9 (-2.4, -1.3) |
| Sex |  |  |  |  |  |  |  |  |
| Male | 48.60 | 40.90 | 30.34 | -2.4 (-2.7, -2.1) | 771.14 | 730.26 | 630.44 | -1.1 (-1.5, -0.7) |
| Female | 41.51 | 33.77 | 28.67 | -2.1 (-2.6, -1.7) | 635.35 | 601.79 | 522.87 | -1.0 (-1.4, -0.6) |
| **Age at death, 1-4 y** |  |  |  |  |  |  |  |  |
| Overall | 1.43 | 1.14 | 0.95 | -2.0 (-2.7, -1.2) | 34.22 | 29.13 | 24.25 | -1.9 (-2.1, -1.7) |
| Race/ethnicity |  |  |  |  |  |  |  |  |
| NH white | 1.14 | 0.96 | 0.68 | -2.4 (-3.6, -1.2) | 30.13 | 25.65 | 22.42 | -1.6 (-1.8, -1.3) |
| NH black | 2.43 | 1.76 | 1.67 | -1.8 (-3.1, -0.4) | 54.59 | 47.44 | 40.15 | -1.5 (-2.1, -0.9) |
| Hispanic | 1.54 | 1.16 | 0.96 | -2.1 (-3.4, -0.8) | 30.87 | 26.52 | 19.40 | -2.8 (-3.5, -2.1) |
| NH other | 1.55 | 1.26 | 1.17 | -1.2 (-3.4, 1.1) | 31.04 | 24.82 | 19.82 | -2.4 (-3.0, -1.7) |
| Sex |  |  |  |  |  |  |  |  |
| Male | 1.63 | 1.21 | 1.03 | -2.4 (-3.1, -1.6) | 37.92 | 31.29 | 27.28 | -1.9 (-2.1, -1.7) |
| Female | 1.23 | 1.07 | 0.87 | -1.5 (-2.7, -0.3) | 30.35 | 26.88 | 21.08 | -1.9 (-2.2, -1.7) |
| **Age at death, 5-17 y** |  |  |  |  |  |  |  |  |
| Overall | 0.46 | 0.3 | 0.28 | -3.0 (-3.6, -2.3) | 27.20 | 23.11 | 19.42 | -1.8 (-2.3, -1.3) |
| Race/ethnicity |  |  |  |  |  |  |  |  |
| NH white | 0.42 | 0.30 | 0.25 | -3.3 (-4.1, -2.5) | 26.14 | 21.99 | 18.61 | -1.8 (-2.4, -1.1) |
| NH black | 0.69 | 0.42 | 0.49 | -2.1 (-3.3, -1.0) | 37.32 | 31.49 | 29.59 | -1.2 (-2.2, -0.2) |
| Hispanic | 0.41 | 0.20 | 0.22 | -2.6 (-4.1, -1.1) | 22.62 | 20.72 | 15.99 | -2.0 (-3.2, -0.8) |
| NH other | 0.38 | 0.23 | 0.38 | -1.4 (-3.6, 1.0) | 21.91 | 18.97 | 15.04 | -1.9 (-3.3, -0.4) |
| Sex |  |  |  |  |  |  |  |  |
| Male | 0.58 | 0.33 | 0.35 | -2.9 (-3.6, -2.1) | 33.48 | 28.91 | 24.23 | -1.7 (-2.6, -0.9) |
| Female | 0.35 | 0.26 | 0.21 | -3.1 (-3.8, -2.3) | 20.57 | 17.03 | 14.41 | -1.8 (-2.6, -1.0) |
| **Age at death, 18-34 y** |  |  |  |  |  |  |  |  |
| Overall | 0.61 | 0.49 | 0.36 | -3.3 (-3.8, -2.8) | 97.15 | 103.7 | 115.03 | 1.1 (0.3, 1.8) |
| Race/ethnicity |  |  |  |  |  |  |  |  |
| NH white | 0.65 | 0.48 | 0.35 | -3.8 (-4.4, -3.2) | 86.99 | 98.90 | 120.84 | 1.9 (1.4, 2.4) |
| NH black | 0.71 | 0.74 | 0.50 | -2.8 (-3.8, -1.8) | 174.00 | 169.41 | 163.95 | -0.2 (-1.0, 0.6) |
| Hispanic | 0.44 | 0.37 | 0.37 | -1.8 (-3.5, 0.0) | 83.36 | 85.98 | 83.10 | 0.1 (-1.3, 1.5) |
| NH other | 0.44 | 0.40 | 0.18 | -1.6 (-3.7, 0.6) | 62.60 | 60.71 | 65.00 | 0.3 (-0.1, 0.8) |
| Sex |  |  |  |  |  |  |  |  |
| Male | 0.67 | 0.66 | 0.49 | -2.6 (-3.3, -1.9) | 136.15 | 148.45 | 160.97 | 1.0 (0.4, 1.6) |
| Female | 0.56 | 0.31 | 0.23 | -4.7 (-5.4, -4.0) | 57.11 | 57.7 | 67.35 | 0.9 (0.6, 1.2) |
| **Age at death, 35-49 y** |  |  |  |  |  |  |  |  |
| Overall | 0.64 | 0.48 | 0.38 | -2.2 (-2.7, -1.7) | 243.12 | 246.08 | 235.24 | -0.2 (-0.5, 0.1) |
| Race/ethnicity |  |  |  |  |  |  |  |  |
| NH white | 0.69 | 0.54 | 0.46 | -1.5 (-2.1, -0.8) | 219.33 | 239.87 | 251.40 | 0.8 (0.5, 1.1) |
| NH black | 0.70 | 0.45 | 0.36 | -2.5 (-3.7, -1.2) | 464.41 | 410.49 | 349.13 | -1.6 (-2.2, -1.0) |
| Hispanic | 0.39 | 0.35 | 0.28 | -2.7 (-4.1, -1.2) | 190.06 | 169.58 | 151.86 | -1.3 (-1.9, -0.6) |
| NH other | 0.32 | 0.25 | 0.12 | -2.4 (-4.6, -0.2) | 140.36 | 136.67 | 128.63 | -0.6 (-1.1, -0.1) |
| Sex |  |  |  |  |  |  |  |  |
| Male | 0.69 | 0.54 | 0.46 | -1.7 (-2.3, -1.1) | 313.10 | 309.14 | 295.93 | -0.3 (-0.6, -0.1) |
| Female | 0.60 | 0.42 | 0.32 | -2.8 (-3.6, -2.0) | 174.40 | 183.98 | 175.21 | 0.0 (-0.3, 0.4) |
| **Age at death, 50-64 y** |  |  |  |  |  |  |  |  |
| Overall | 0.65 | 0.46 | 0.4 | -2.4 (-3.4, -1.4) | 795.63 | 737.44 | 751.54 | -0.3 (-0.4, -0.2) |
| Race/ethnicity |  |  |  |  |  |  |  |  |
| NH white | 0.67 | 0.47 | 0.45 | -1.9 (-3.1, -0.8) | 753.99 | 714.15 | 762.83 | 0.1 (-0.1, 0.3) |
| NH black | 0.74 | 0.70 | 0.37 | -3.5 (-5.6, -1.3) | 1368.62 | 1198.37 | 1099.00 | -1.2 (-1.4, -1.0) |
| Hispanic | 0.38 | 0.31 | 0.24 | -2.8 (-4.9, -0.7) | 596.78 | 537.36 | 496.07 | -1.1 (-1.2, -0.9) |
| NH other | 0.47 | 0.19 | 0.20 | -3.4 (-6.3, -0.3) | 474.91 | 411.44 | 407.04 | -0.7 (-0.9, -0.5) |
| Sex |  |  |  |  |  |  |  |  |
| Male | 0.72 | 0.53 | 0.46 | -1.4 (-2.2, -0.5) | 991.35 | 923.47 | 938.71 | -0.3 (-0.4, -0.2) |
| Female | 0.59 | 0.41 | 0.35 | -3.0 (-3.7, -2.2) | 612.43 | 562.25 | 574.93 | -0.3 (-0.5, -0.2) |
| **Age at death, ≥65 y** |  |  |  |  |  |  |  |  |
| Overall | 1.45 | 0.83 | 0.64 | -4.2 (-4.9, -3.5) | 5165.06 | 4734.20 | 4065.00 | -1.5 (-1.6, -1.4) |
| Race/ethnicity |  |  |  |  |  |  |  |  |
| NH white | 1.50 | 0.87 | 0.69 | -3.9 (-4.6, -3.2) | 5257.74 | 4909.71 | 4285.54 | -1.3 (-1.4, -1.2) |
| NH black | 1.39 | 0.86 | 0.43 | -5.0 (-6.8, -3.2) | 5771.03 | 5042.22 | 4203.58 | -1.8 (-2.0, -1.5) |
| Hispanic | 1.15 | 0.30 | 0.50 | -5.4 (-9.1, -1.6) | 3464.59 | 3148.16 | 2817.09 | -1.2 (-1.4, -1.0) |
| NH other | 0.42 | 0.65 | 0.52 | -4.1 (-6.4, -1.7) | 2915.30 | 2635.32 | 2350.02 | -1.2 (-1.3, -1.1) |
| Sex |  |  |  |  |  |  |  |  |
| Male | 1.27 | 0.81 | 0.63 | -3.7 (-5.2, -2.2) | 5635.32 | 5002.52 | 4322.99 | -1.5 (-1.7, -1.3) |
| Female | 1.59 | 0.84 | 0.65 | -4.5 (-5.7, -3.2) | 4836.98 | 4537.22 | 3859.25 | -1.3 (-1.5, -1.0) |

Footnote: AAPC, average annual percent change; CI, confidence interval
Mortality rates are calculated as the number of deaths per 100,000 population for all age groups one year or older and as the number of deaths per 100,000 live births for the infant (<1 year) age group.
Mortality rates for "all ages" are age-adjusted; mortality rates for individual age groups are crude (age-specific).

All statistical tests were performed with SAS version 9.4 (Cary, NC) and the Joinpoint Regression Program version 4.7.0.0.^25^

## RESULTS

### All cause and age-adjusted CHD mortality

#### Overall all-cause and CHD mortality

In the United States between 1999 and 2017, there were approximately 2.5 million deaths annually with 3,084 per year due to CHD, representing 1 in 814 deaths. While the all-cause, age-adjusted mortality rate decreased on average by 1.1% per year (Table 1), the mean annual decrease in mortality due to CHD and CHD-related mortality was faster, at 2.6% and 2.0%, respectively. Among all CHD-related deaths, CHD was listed as the UCOD 65.2% of the time (Supplemental Table 2).

**Table 2.** Temporal Trends in Infant Mortality Resulting From CHD, by CHD lesion, 1999–2017

| Defect | Deaths in all age groups | Deaths during infancy | % deaths that occur during infancy | 99-17 AAPC in infant mortality (95% CI) | Median Age at Death 1999-2006 | Median Age at Death 2007-2017 |
| --- | --- | --- | --- | --- | --- | --- |
| Anomalous pulmonary venous connection | 661 | 591 | 89.4 | -1.0 (-2.8, 0.9) | 30 days | 30 days |
| Aortic valve anomalies | 946 | 449 | 47.5 | -4.9 (-6.5, -3.2) | 152 days | 23 years |
| Atrioventricular septal defect | 1033 | 606 | 58.7 | 0.6 (-2.5, 3.8) | 183 days | 213 days |
| Coarctation of the aorta | 898 | 517 | 57.6 | -3.2 (-5.0, -1.3) | 61 days | 152 days |
| Common truncus | 796 | 619 | 77.8 | -1.9 (-5.1, 1.4) | 30 days | 30 days |
| Common ventricle/double inlet ventricle | 482 | 260 | 53.9 | -3.5 (-5.2, -1.7) | 183 days | 335 days |
| Ebstein anomaly | 850 | 485 | 57.1 | -1.7 (-2.7, -0.6) | 30 days | 91 days |
| Hypoplastic left heart syndrome | 6536 | 5626 | 86.1 | -2.8 (-3.6, -1.9) | 14 days | 30 days |
| Pulmonary artery atresia/stenosis | 1493 | 855 | 57.3 | -1.6 (-2.9, -0.3) | 183 days | 213 days |
| Pulmonary valve anomalies | 172 | 128 | 74.4 | 0.7 (-2.7, 4.2) | 30 days | 61 days |
| Tetralogy of Fallot | 3103 | 1176 | 37.9 | -0.9 (-3.2, 1.5) | 8 years | 16 years |
| Transposition of the great arteries | 2165 | 1243 | 57.4 | -3.1 (-4.3, -2.0) | 61 days | 213 days |
| Tricuspid valve anomalies | 439 | 165 | 37.6 | -2.2 (-5.7, 1.5) | 12 years | 17 years |
| Ventricular septal defect | 2613 | 592 | 22.7 | -5.6 (-7.5, -3.7) | 44 years | 51 years |
| Other specified congenital heart disease | 10900 | 2311 | 21.2 | -2.0 (-2.8, -1.2) | 36 years | 39 years |
| Unspecified congenital heart disease | 20663 | 11134 | 53.9 | -1.9 (-2.3, -1.4) | 213 days | 244 days |

Footnote: AAPC, average annual percent change; CI, confidence interval
Only deaths in which the CHD is listed as the underlying cause of death are included in this table.
Median age at death is calculated among all cases, not just deaths during infancy. Expressed in days when less than 1 year, otherwise expressed in years.

#### All-cause and CHD mortality by sex

Males, compared to females, experienced higher all-cause, age-adjusted mortality (925.5 vs. 661.1 per 100,000) and age-adjusted mortality due to CHD (1.13 vs. 0.93 per 100,000). When evaluating temporal trends, age-adjusted mortality decreased from 1999 and 2017 for both males and females, and this decline was more pronounced for mortality due to CHD than for all-cause mortality (Table 1). Trends in age-adjusted CHD mortality were similar for males and females, with a 3-4% annual decrease between 1999 and 2009, followed by a slower annual decrease of 1.4% through 2017 (Figure 1).

**Figure 1.**
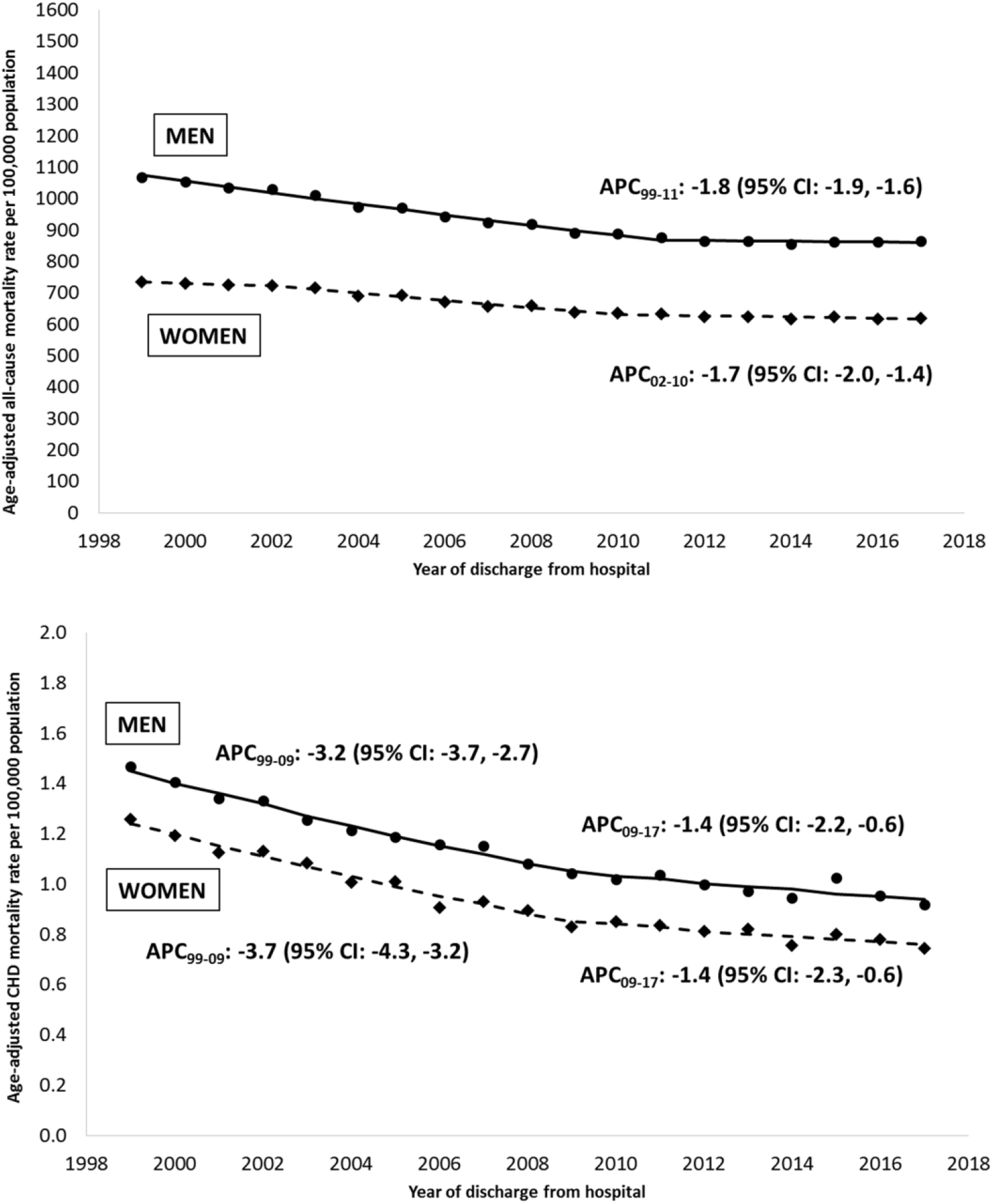
Temporal trends in age-adjusted all-cause mortality (top) and CHD-specific mortality (bottom), by sex, 1999-2017. Footnotes: APC, annual percent change; CHD, congenital heart defect Only APCs that reflect a statistically significant temporal trend in mortality are presented as text.

#### All-cause and CHD mortality by race/ethnicity

Age-adjusted mortality decline from 1999 to 2017 was more pronounced for mortality due to CHD than for all-cause mortality for all race/ethnicities (Figure 2). Age-adjusted all-cause mortality demonstrated that non-Hispanic Blacks experienced more pronounced decreases in rates of mortality (from 1149.3 to 882.2 per 100,000) than non-Hispanic Whites (from 859.6 to 755.1 per 100,000) and Hispanics (676.2 to 525.6 per 100,000, Table 1). In contrast, non-Hispanic blacks experienced a similar rate of decline in CHD-specific age-adjusted mortality at 2.3% annually, compared to 2.6% for non-Hispanic whites (Figure 2). Thus, whereas the age-adjusted mortality rate ratio comparing non-Hispanic blacks to whites has decreased from 1.34 to 1.17 for all-cause mortality, the disparity for CHD-specific mortality has actually risen from 1.24 in 1999 to 1.31 in 2017 without a consistent temporal trend (Figure 3). Among Hispanics, although decreases in age-adjusted CHD mortality mirrored non-Hispanic whites from 1999-2009, since then the static rate among Hispanics and the ongoing decline among non-Hispanic whites has resulted in 2017 being the first time during the 19-year study period that CHD mortality is higher for Hispanics than whites (Figure 3).

**Figure 2.**
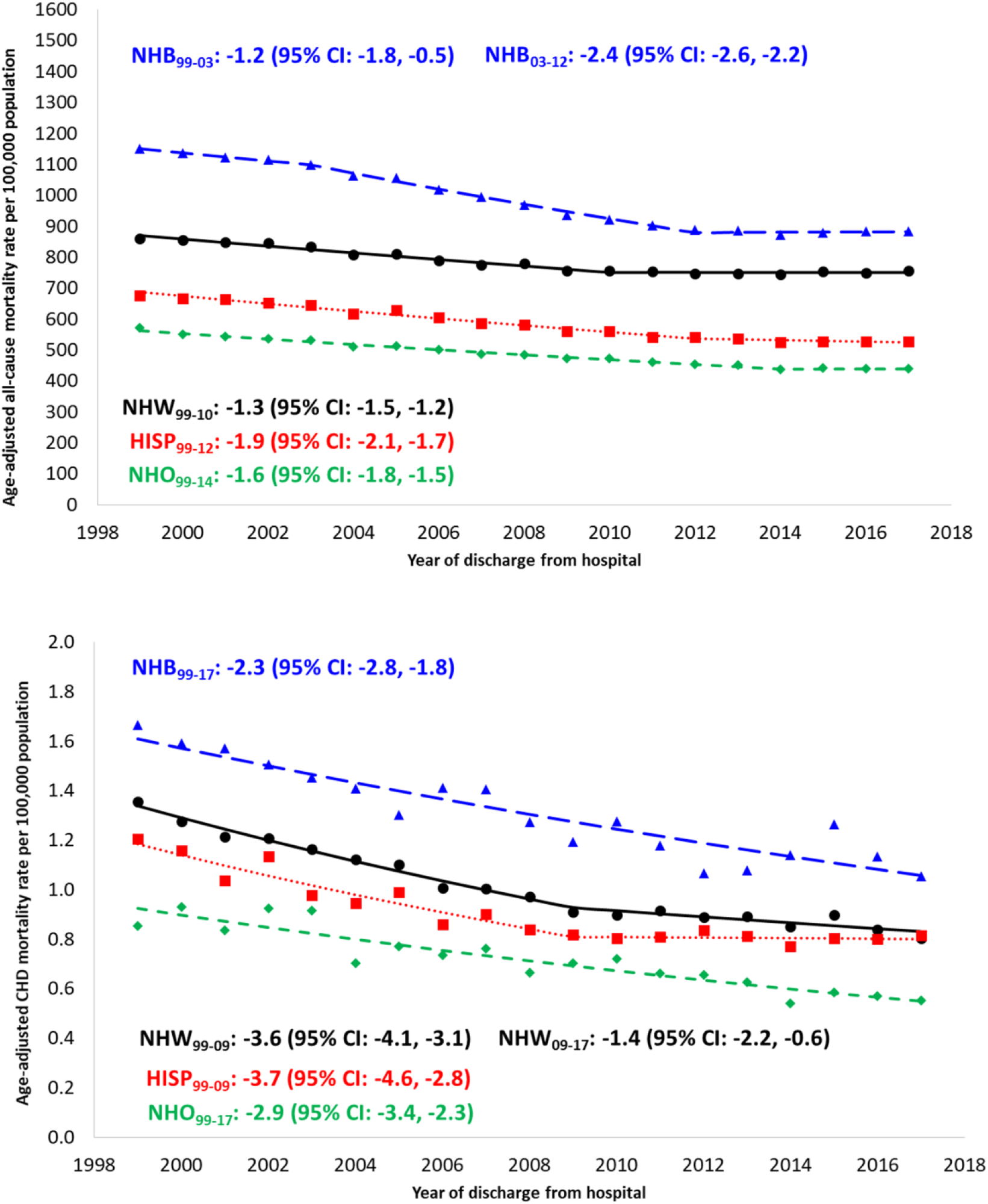
Temporal trends in age-adjusted all-cause mortality (top) and CHD-specific mortality (bottom), by race/ethnicity, 1999-2017. Footnotes: APC, annual percent change; CHD, congenital heart defect Only APCs that reflect a statistically significant temporal trend in mortality are presented as text.

**Figure 3.**
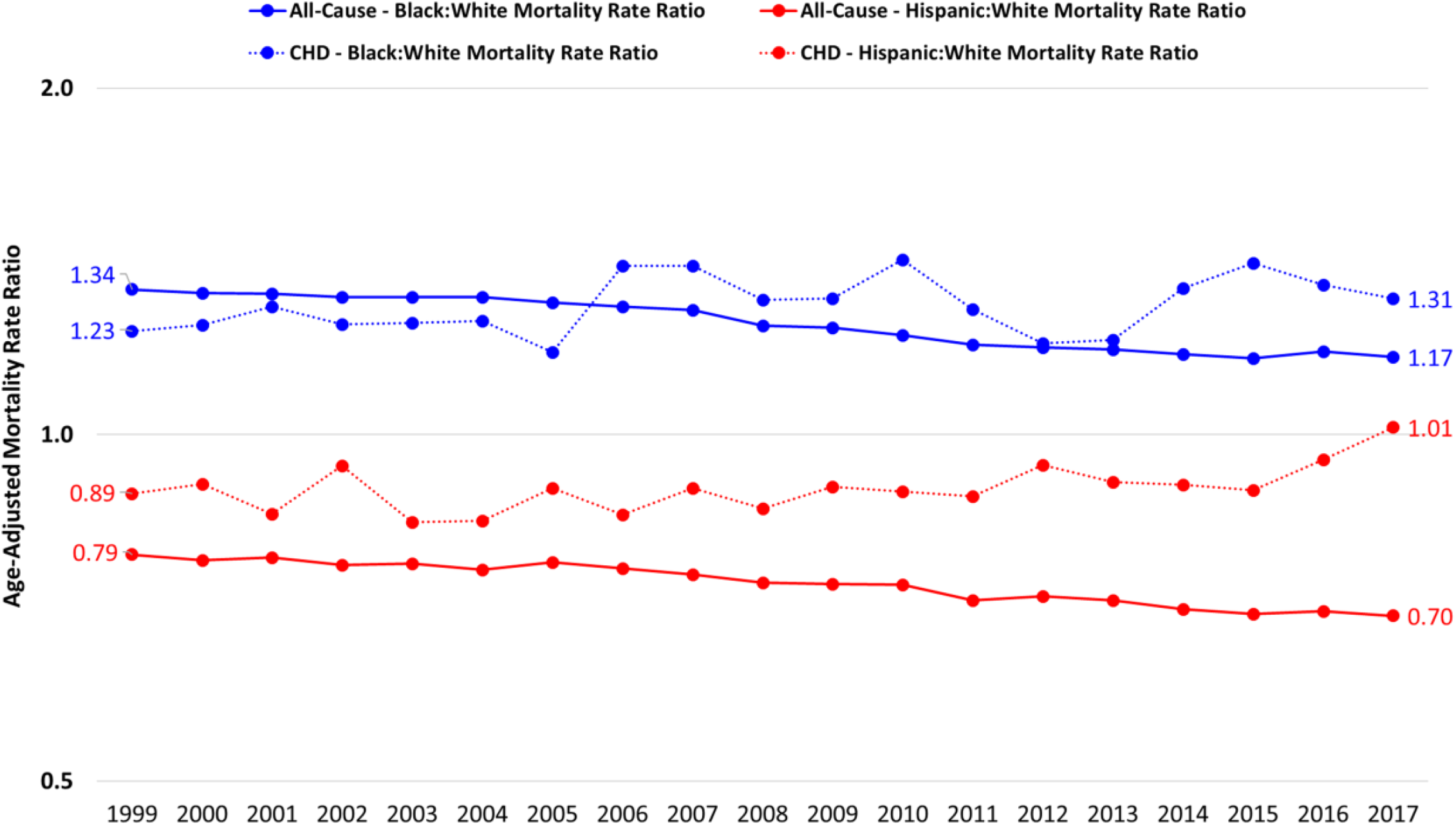
Age-standardized mortality rate ratios, all ages, 1999-2017. Footnotes: Rate ratios were calculated as the age-standadrized mortality rate in one racial/ethnic group (e.g., non-Hispanic blacks) divided by the age-standadrized mortality rate in the reference racial/ethnic group (e.g., non-Hispanic whites). This was done deparately for all-cause mortality and mortality due to CHD.

#### CHD mortality by CHD lesion

Overall, the largest number of deaths in which a CHD was listed as the UCOD was “other specified congenital heart disease” and “unspecified congenital heart disease”. Over the 19-year period, the lesions with the highest percentage of death that occurred during infancy were: anomalous pulmonary venous connection (89.4%), hypoplastic left heart syndrome (86.1%) and common truncus (e.g. truncus arteriosus, 77.8%) (Table 2) When examining temporal trends, infant mortality due to CHD had a statistically significant decrease for most CHD lesions (Table 2). The highest mean annual decrease in infant mortality by lesion over the 19-year period was for ventricular septal defect (5.6%), aortic valve anomalies (4.9%) and common ventricle/double inlet left ventricle (3.5%). For most CHD lesions, we saw an increase in the median age at death between the early (1999-2006) and late (2007-2017) time periods. Of note, for the lesions associated with the highest infant mortality, anomalous pulmonary venous connection and common truncus, the median age at death did not change between these two time periods, and remained at 30 days. For hypoplastic left heart syndrome, the median age at death increased by 53% (from 14 days to 30 days).

### Age-specific CHD mortality

#### Infant mortality

Death in infancy accounted for 47.7% (n=27,962) of all deaths due to CHD. Infant mortality was consistently higher in males than in females throughout the study period though decreases in mortality over time were similar between the sexes (Table 1). Although all race/ethnic groups experienced overall reductions in CHD-specific infant mortality, non-Hispanics Black infants experienced the highest mortality due to CHD, followed by Hispanics (Table 1, Figure 5). The annual percent decrease during the study period for non-Hispanic blacks was less (1.8%) than the decrease among non-Hispanic whites (2.6%). Hispanics, despite experiencing the most significant improvements from 1999-2004, have had unchanged CHD-specific infant mortality for over a decade (Figure 5). These differences in temporal trends have resulted in mortality rate ratios for non-Hispanic blacks and Hispanics that reflect worsening disparities in CHD-specific infant mortality (Figure 6).

**Figure 5.**
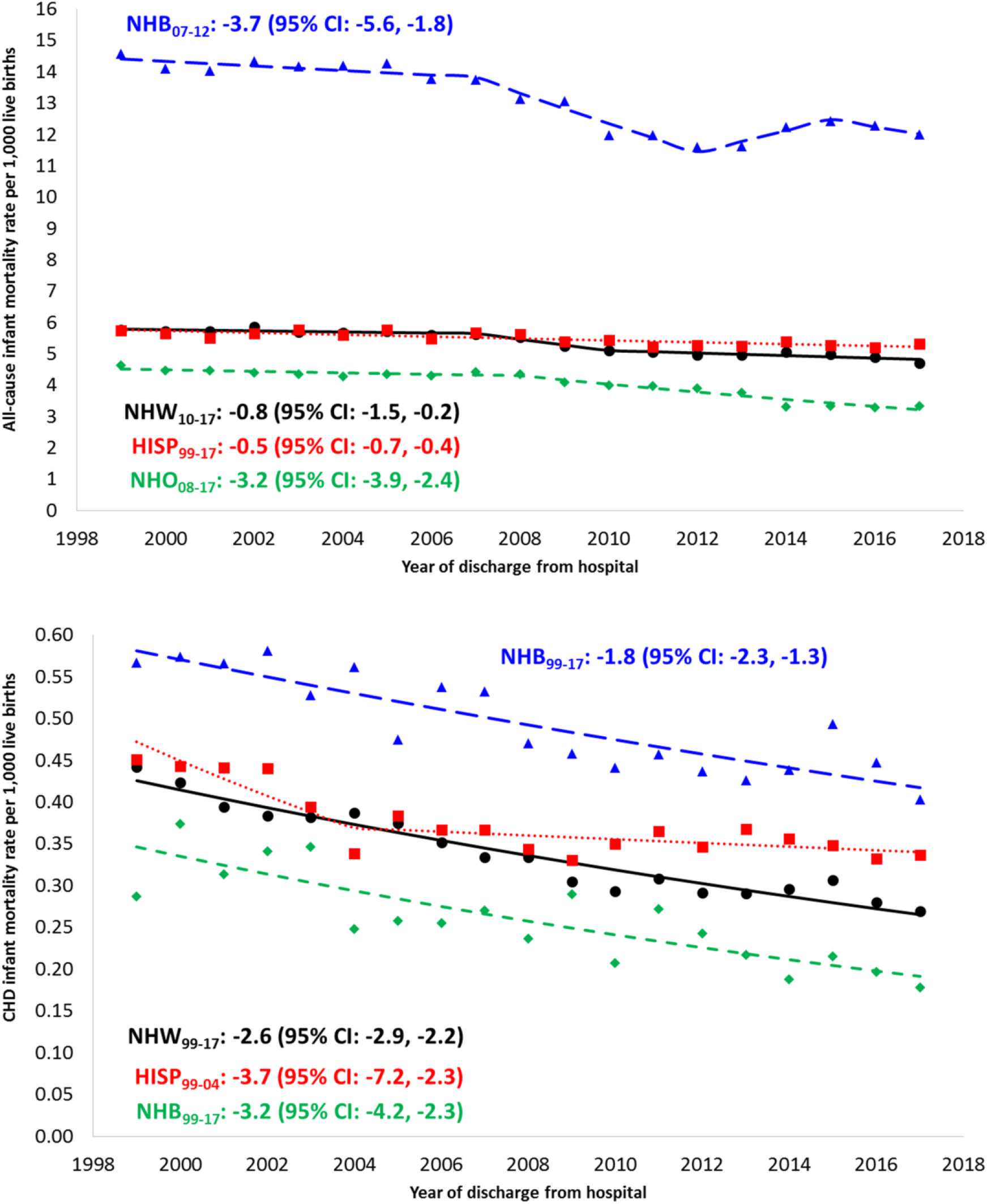
Temporal trends in all-cause infant mortality (top) and CHD-specific infant mortality (bottom), by race/ethnicity, 1999-2017. Footnotes: APC, annual percent change; CHD, congenital heart defect Only APCs that reflect a statistically significant temporal trend in mortality are presented as text.

**Figure 6.**
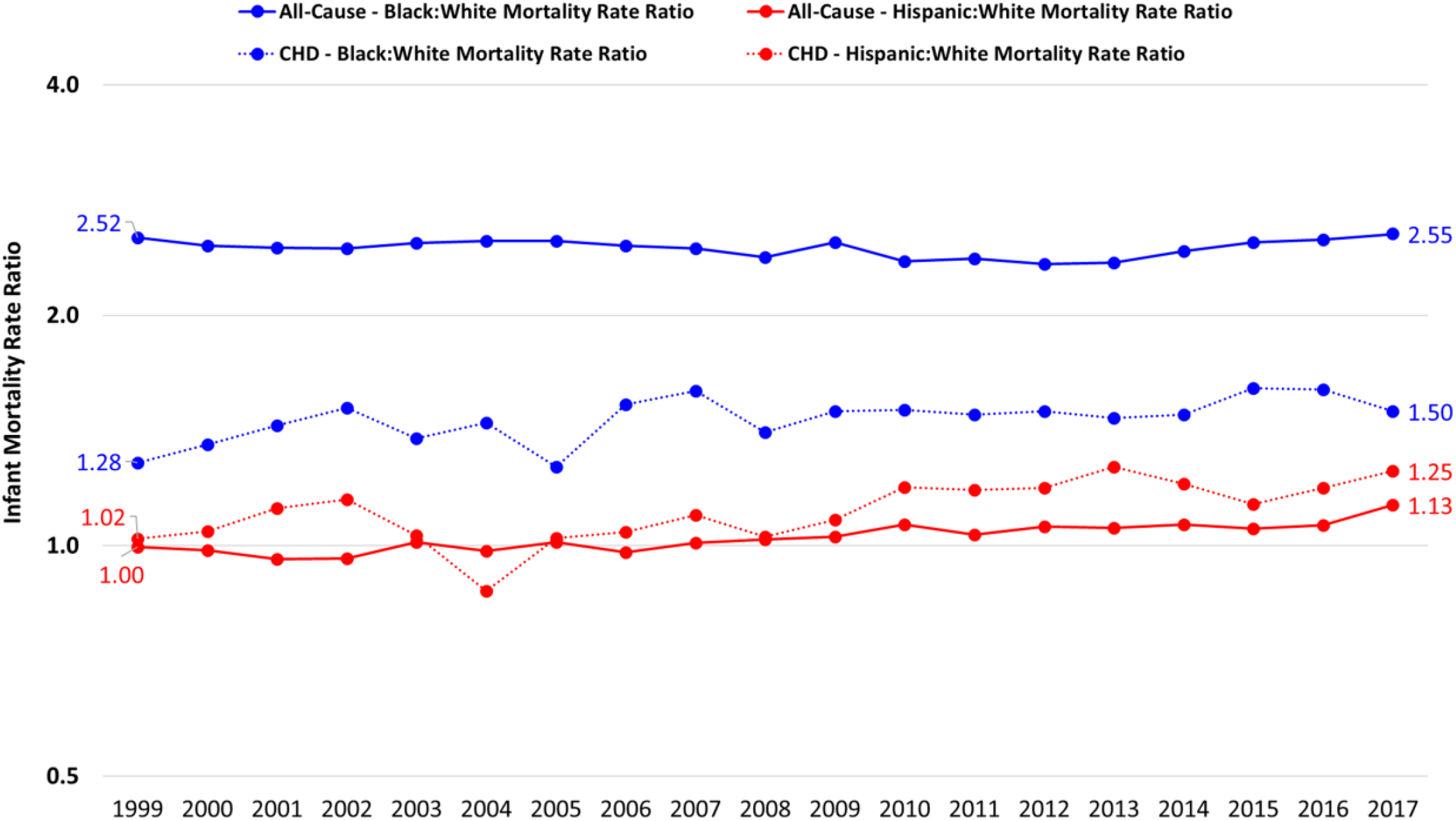
Infant mortality rate ratios, all ages, 1999-2017. Footnotes: Rate ratios were calculated as the infant mortality rate in one racial/ethnic group (e.g., non-Hispanic blacks) divided by the infant mortality rate in the reference racial/ethnic group (e.g., non-Hispanic whites). This was done deparately for all-cause infant mortality and infant mortality due to CHD.

#### Childhood mortality

For those age 1-17 years, mortality due to CHD decreased between 1999 and 2017. Among children aged 1 to 4 years mortality decreased from 1.43 to 0.95 per 100,000, with similar findings in higher rates of mortality for males and for Blacks and Hispanics. Among children 5-17 years old, the age group with the lowest CHD-specific mortality rates decreased from 0.46 to 0.28 per 100,000. While there were higher rates of mortality for males and for Blacks, there were lower mortality rates for Hispanics (Supplemental Table 2). It was during childhood when the highest proportion of CHD-related deaths had the CHD listed as the UCOD (Supplemental Table 2).

#### Adult mortality

Adult mortality resulting from CHD decreased among all adults during the study period, with the most pronounced decreases in the oldest (>65 years) age group (Table 1). In younger adults (18-34 years), non-Hispanic Whites experienced the most significant decreases over time, whereas non-Hispanic Blacks and Hispanics tended to have higher-magnitude decreases among older adults (Table 1).

#### Comparing all age-specific CHD mortality

Unlike the characteristic J-shaped association observed between age and all-cause mortality, mortality due to CHD was at least 30-fold higher during infancy than any other age group (Figure 1). CHD mortality decreased through the 5-17 year old age group, followed by a subsequent increase with older age groups. The largest decrease in mortality due to CHD was among individuals ≥ 65 years (-4.2% each year); whereas in 1999 they had the second-highest mortality of any age group (1.5 per 100,000) and comparable with early childhood CHD mortality (1.4 per 100,000), by 2017, mortality in this oldest age group was 40% lower than among children aged 1-4 years (Figure 4). It is noteworthy that all seven age groups experienced at least a 2% average annual decrease in mortality due to CHD.

**Figure 4.**
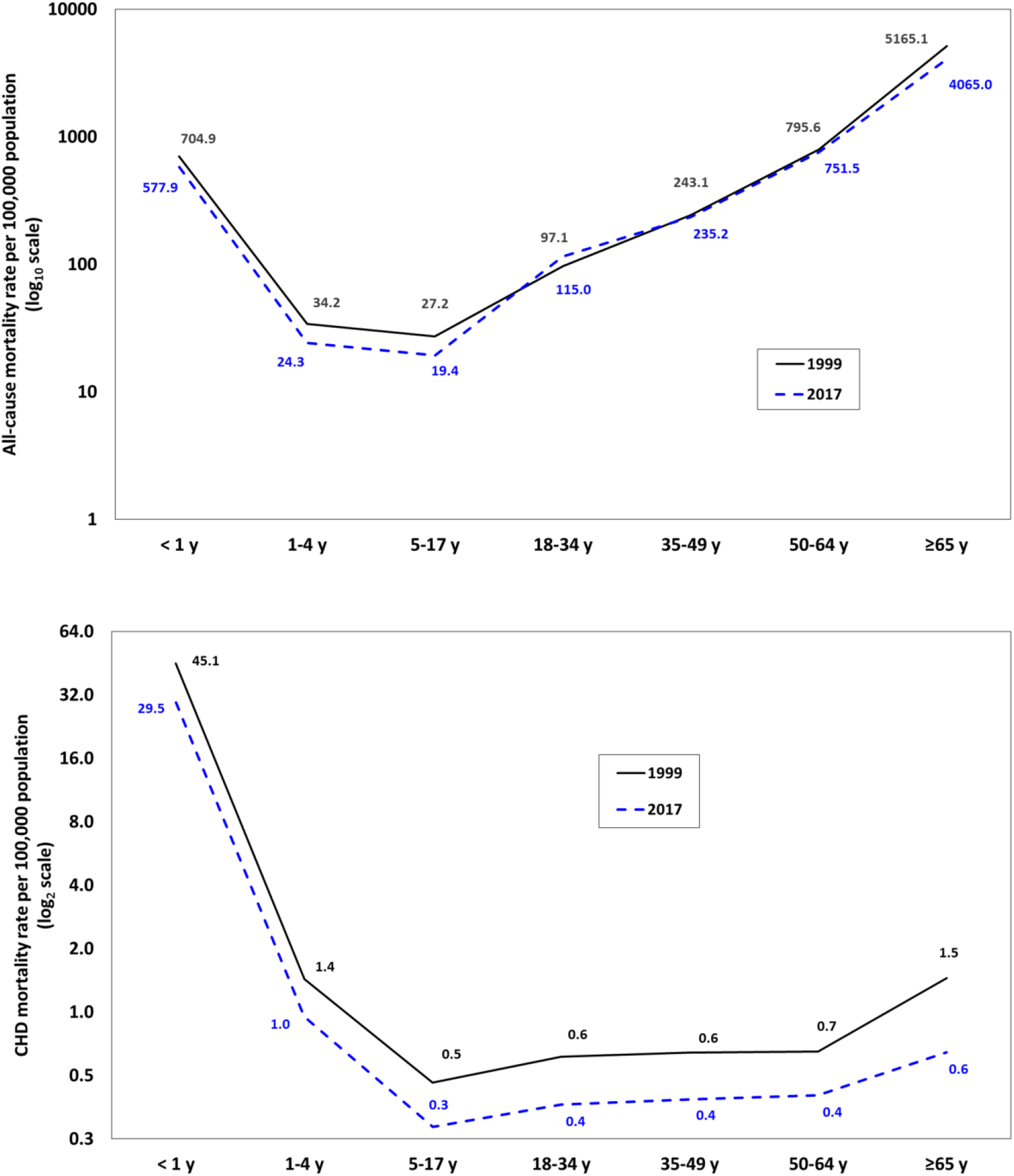
All-cause mortality (top) and CHD-specific mortality (bottom), by age group, 1999 and 2017. Footnotes: Due to the large differences in the magnitude of all-cause versus CHD-specific mortality rates, to visualize and compare their respective distributions by age, all-cause mortality rates were plotted on the Y-axis with a log base 10 transformation (common logarithm), whereas CHD-specific mortality rates were plotted on the Y-axis with a log base 2 transformation (binary logarithm).

#### Sensitivity analyses

Among all age groups combined, 7,169 (12.3%) of the 58,599 deaths due to CHD had prematurity (infants only), an extracardiac defect, or a genetic syndrome listed as a contributing cause of death (Supplemental Table 1). We found no appreciable differences in comparison of rates by decedent characteristics or across time when these cases were excluded from the numerator. However, among infants, nearly 19% of deaths due to CHD had one of these conditions listed as a contributing cause. When these cases were excluded from the numerator, the annual percent decrease in CHD-specific infant mortality was mildly more pronounced (-2.9; 95% CI: -3.1, -2.6) when these cases were excluded than when they were included (-2.2; 95% CI: -2.6, -1.8; p=0.047).

## DISCUSSION

This work provides a contemporary update to previous studies on CHD mortality by age, sex and race/ethnicity in the United States.^5,6,26,27^ Overall during the 19-year study period, CHD mortality decreased 39.4%, a more pronounced change relative to the 24% drop that was observed in the prior study’s timeframe (1999-2006). Nearly half of all CHD mortality (47.7%) occurred during infancy, which has decreased only (0.4%) compared to the prior study’s time frame (1999-2006). This may be in part due to lack of several novel surgical techniques and more surgeries in premature infants in the most recent decades. Nearly one quarter of all CHD mortality (23.5%) occurred among those >18 years for the entire cohort, and the median age at death for most CHD lesions increased with time, likely due to advances in surgical and medical care.

Since the last year of the prior national assessment of CHD mortality, overall CHD mortality rates for males and females have decreased. However, when the average annual percent change in morality is compared between the two groups, the difference between males and females has not changed significantly in the last 19 years (Figure 1). This was notably true throughout the lifespan, from birth to those >65years of age. This is different than other studies that have shown an increased mortality for females in infancy and childhood. At least two large studies have showed that for infants, there is an increased risk of death in early infancy for girls after high-risk cardiac operations ^**28**,**29**^ Another study showed that among children surviving their initial congenital heart surgery, there was an increased risk of death from CHD in females^**30**^. This is in contrast to some studies on adults with CHD, one of the largest of which did not show differences in mortality between men and women.^**31**^ Some differences may be due to prevalence of different CHDs in different sexes as well as certain higher risk lesions being present in specific sexes^**32**^. For example, female infants have lower rates of severe CHD than males (thus contributing to lower CHD-related mortality as a proportion of all-cause mortality), but higher mortality in those that do have severe CHD requiring high-risk procedures.^**28**,**29**^Currently, little is still known about the underlying mechanisms of sex differences in outcomes, and thus genetic, hormonal, behavioral or other factors need to be taken into account. ^**32**^ Given our findings, sex should at the very least be considered in effect modification of certain conditions as well as in risk stratification in conditions in which there is evidence of sex-related increased risk of morbidity and mortality.^**32**^

Our study describes the most current nationwide data on racial/ethnic mortality disparities in CHD populations^**5**,**6**^. Our study demonstrates that even when removing the impact of CHD deaths for which either prematurity or extracardiac birth defects are a contributing cause, there remains a racial disparity most pronounced between non-Hispanic Blacks and non-Hispanic Whites in the first year of life. Disconcertingly, this Black:White disparity in CHD mortality persists through about 35 years of age. Also concerning is the novel finding that Hispanics went from being 11% less likely to die from CHD in 1999 to having the same mortality rate as non-Hispanic Whites in 2017. Determination of the multifactorial nature of these finding may help to reduce disparities.

Racial/ethnic disparities and associated difference in mortality rates are not isolated to the individuals affected by CHD. Children of different racial and ethnic backgrounds have increased mortality in other congenital defects and chronic diseases^**33**^. One study showed that US children of non-Hispanic Black and Hispanic mothers had significantly higher risks of mortality in children less than 8 years old for 1/3-1/2 of 21 analyzed birth defects.^**26**^ Interestingly, five of these defects were of cardiac origin for non-Hispanic black children and four for Hispanic children. Thus, the persistence in the racial/ethnic disparities in mortality despite the improvement in overall CHD (and other chronic disease) mortality make clear that there are contributions at several other levels-population, systemic, institutional, and individual -that facilitate its existence.

At the population level, access to quality care can be an issue. Families living in rural communities have a higher risk of missing a prenatal congenital heart disease diagnosis.^**34**^ Furthermore, CHD care is typically concentrated at specialized urban pediatric cardiac centers given that the expertise and resources needed to manage complex patients. A recent study demonstrated that lack of proximity to one of these top 50 centers resulted in an infant mortality rate that was 28% greater than for infants whose mothers lived proximal to a center. ^**35**^

At the systemic level, national health policies that protect the insurance status of patients with CHD including non-exclusion due to pre-existing conditions or lifetime caps are critical to reducing racial/ethnic disparities throughout the lifespan (ref?). Furthermore, as there are increasing numbers of individuals with CHD surviving into adulthood, assuring access to high quality inpatient and outpatient adult CHD care and increasing numbers of adult CHD providers is critical to the long term reduction in racial/ethnic disparities in CHD mortality.^36,37^

At an institutional level, expertise in CHD surgery, surgical volume, and CHD perioperative care are important factors to reducing racial/ethnic disparities, as there may be referral patterns for lower socioeconomic status patients to “low performing” hospitals.^38–40^ One multicenter database study demonstrated Hispanic ethnicity was associated with increased odds of experiencing a CHD-related complication, and blacks, compared to whites, had higher rates of mortality after experiencing a CHD-related complication.^41^ Additionally, implicit bias may play a role in racial/ethnic disparities. One multicenter study demonstrated that after adjusting for age, sex, genetic syndrome, surgery risk category, insurance type and hospital of surgery, the relative risk of death was 1.32 for non-Hispanic Blacks (CI, 1.14-1.52) and 1.21 for Hispanics (CI, 1.07-1.37), both compared with non-Hispanic whites.^42^ Thus understanding referral patterns, CHD center quality, and implementing implicit bias training are all pieces necessary to reduce racial/ethnic disparities in mortality in the CHD.

At the individual level, while earlier studies showed that maternal race/ethnicity was a predictor of increased risk of mortality^8^, several recent studies have demonstrated that there are various socioeconomic mediators that play a role in the differences seen in racial/ethnic mortality and CHD. One study showed that maternal education and insurance status explained 33.2% and 27.6% of the relationship between race/ethnicity and poor outcome.^43^ Another recent study demonstrated that adjusting for insurance type reduced the Black-White disparity in CHD mortality risk by 50%. ^44^ Transportation, opportunity costs and knowledge of the need for ongoing outpatient care may also play a role in these racial/ethnic disparities. One study showed that minorities with CHD have an elevated risk of lapse in care in outpatient clinical follow up.^45^ Other studies have shown that many of the racial/ethnic differences in mortality were in the outpatient setting. One statewide study noted that racial/ethnic differences in mortality were most notably observed during the postneonatal period and early childhood.^46^ A more recent study demonstrated that once children with CHD are released from the hospital, those from low-income neighborhoods have a higher risk of mortality and utilize more inpatient resources as compared to higher income communities.^47^ Thus, putting into place programs that can assist with maintenance long-term outpatient care and patient/parent education about the need for lifelong care plays a major role in reducing racial/ethnic disparities.

Unfortunately, the much of mortality due to CHD is documented as unspecified congenital heart disease (35%). This is lack of specificity even greater during infancy (40%). This makes it challenging to draw reliable conclusions about the impact of specific CHD lesions over time. That being said, we consistently see declines in CHD lesion specific infant mortality in the 19 year period, and the median age of death is also consistently higher in more recent years.

As with any study there are limitations when using death certificate data, particularly as it relates to CHD mortality; therefore, interpretation of these results should be done with caution.^26,48^ First, previous studies have shown poor (∼50%) sensitivity for recording more common cardiac diseases states, which may be related to the inadequate training of physicians in the completion of death certificates.^48^ One recent study from 2017 assessed errors in the cause and manner of death on death certificates completed by non–Medical Examiners, and found that 51% of death certificates had major errors regardless of whether or not they were physicians.^49^ Another study in Australia investigated the sensitivity and specificity of the national mortality codes in identifying cardiovascular disease (CVD) deaths and found that national mortality coding underestimated the true proportion of CHD and stroke deaths in the cohort by 13.6% and 50.8%, respectively.^50^ While using UCOD has these limitations, this is similar methodology as used in many analyses, including the prior one by Gilboa et al.^5^

In our study, the most commonly listed cause of a CHD-related death from birth to 64 years was “unspecified CHD”, and 33% of all mortality in which a CHD was the UCOD was coded as unspecified CHD, making it challenging to associate specific diseases as the cause of death. While there are rules surrounding “acceptable” causal relationships between cause-of-death codes to ultimately select the correct UCOD, the data are imperfect and are limited by what was recorded on the death certificate itself. ICD-9 and ICD-10 codes have remained an imperfect tool to accurately classify CHD, which is a challenge as it is part of the classification of the MCOD data. One study from 2018 evaluated accuracy of ICD-9 coding versus medical records for adults with congenital heart disease, and found misclassifications occurred in 23% of patients, and that the overall accuracy of a CHD diagnosis was 48.7%.^51^ While ICD-10 has added more categories for CHD, than listed in ICD-9 and thus increased specificity for a number of phenotypes, the classification of CHD causes of death in MCOD data is still a tool for classifying CHD mortality that was not designed with rigorous etiologic research in mind. While other studies have been able to glean more information from the medical record for cause of death, these would not be possible with the MCOD data alone. Additionally, the death certificate data do not allow for assessment of other factors, including late presentations of disease, early or late postoperative deaths, or where the care was received. These factors all contribute to CHD mortality.

While overall US mortality due to CHD has decreased over the last 19 years, disparities in mortality persist for males compared to females and for non-Hispanic Blacks compared to non-Hispanic Whites. Patient sex should at the very least be considered in the risk stratification in conditions in which there is evidence of sex-related increased risk of morbidity and mortality. Racial disparities in CHD mortality continue to persist in the current era and are most notable between infancy and those aged 35 years. This persistence in, and sometimes exacerbation of disparities despite the improvement in overall CHD mortality in all race/ethnic groups makes clear that there are other contributing factors may be playing a role, including contributions at the population, systemic, institutional, and individual levels. Moving forward, it will be important to take these factors into account to be able to mitigate the gap in sex differences and racial disparities in patients with CHD.

## Data Availability

Data available through National Center for Vital statistics and CDC Wonder

## ACKNOWLEDGEMENTS

None

## Sources of Funding

This project was supported by grant number K23 HL127164 (principal investigator: KNL) and grant number K23 HL127266 (principal investigator: SAM) from the National Institutes of Health/National Heart Lung and Blood Institute. The content is solely the responsibility of the authors and does not necessarily represent the official views of the National Institutes of Health.

## Disclosures

The authors have no financial relationships relevant to this article to disclose.

## Figure Legends

**Supplemental Table 1.**
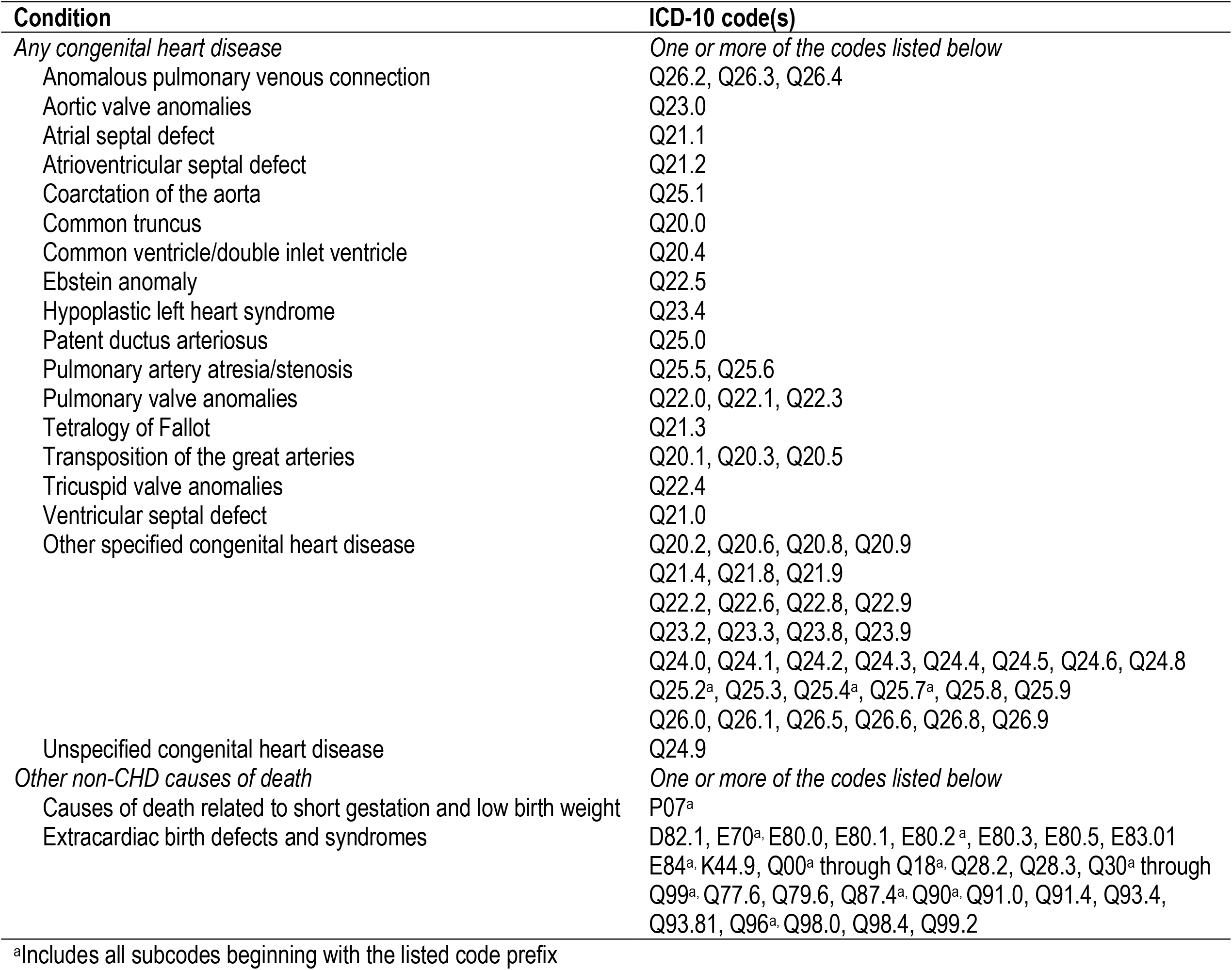
International Classification of Diseases, Tenth Revision (ICD-10) codes used to identify congenital heart disease from Multiple Cause of Death data files, National Center for Health Statistics, United States, 1999-2017

**Supplemental Table 2.** Temporal Trends in Mortality Resulting From CHD and CHD-Related Mortality, by Age, Race/Ethnicity, and Sex, United States, 1999–2017

| Population subgroup | CHD listed as underlying cause of death |  |  |  | CHD listed as underlying or contributing cause of death |  |  |  | % of all CHD-related deaths in which CHD was underlying cause |
| --- | --- | --- | --- | --- | --- | --- | --- | --- | --- |
|  | 1999 | 2006 | 2017 | 99-17 AAPC (95% CI) | 1999 | 2006 | 2017 | 99-17 AAPC (95% CI) |  |
| **All ages** |  |  |  |  |  |  |  |  |  |
| Overall | 1.37 | 1.03 | 0.83 | -2.6 (-2.9, -2.2) | 1.98 | 1.57 | 1.35 | -2.0 (-2.4, -1.6)* | 65.2 |
| Race/ethnicity |  |  |  |  |  |  |  |  |  |
| NH white | 1.35 | 1.01 | 0.80 | -2.6 (-3.0, -2.2) | 1.94 | 1.51 | 1.32 | -2.0 (-2.4, -1.6)* | 64.5 |
| NH black | 1.67 | 1.41 | 1.05 | -2.3 (-2.8, -1.8) | 2.44 | 2.12 | 1.66 | -2.0 (-2.5, -1.6)* | 67.0 |
| Hispanic | 1.20 | 0.86 | 0.81 | -2.2 (-2.8, -1.5) | 1.73 | 1.36 | 1.28 | -1.6 (-2.1, -1.0)* | 65.8 |
| NH other | 0.85 | 0.73 | 0.55 | -2.9 (-3.4, -2.3) | 1.42 | 1.11 | 0.86 | -2.7 (-3.2, -2.2)* | 66.3 |
| Sex |  |  |  |  |  |  |  |  |  |
| Male | 1.47 | 1.16 | 0.92 | -2.4 (-2.8, -2.0) | 2.10 | 1.72 | 1.49 | -1.7 (-2.1, -1.3)* | 66.1 |
| Female | 1.26 | 0.91 | 0.74 | -2.7 (-3.2, -2.3) | 1.87 | 1.41 | 1.21 | -2.2 (-2.7, -1.7)* | 64.1 |
| **Age at death, <1 y** |  |  |  |  |  |  |  |  |  |
| Overall | 45.14 | 37.42 | 29.52 | -2.2 (-2.6, -1.8) | 67.21 | 56.75 | 47.58 | -1.7 (-2.2, -1.3)* | 65.3 |
| Race/ethnicity |  |  |  |  |  |  |  |  |  |
| NH white | 44.18 | 35.12 | 26.89 | -2.6 (-2.9, -2.2) | 64.12 | 51.75 | 42.29 | -2.1 (-2.5, -1.6)* | 66.6 |
| NH black | 56.70 | 53.77 | 40.26 | -1.8 (-2.3, -1.3) | 87.42 | 83.25 | 67.88 | -1.7 (-2.2, -1.2)* | 63.7 |
| Hispanic | 45.08 | 36.62 | 33.66 | -1.8 (-2.6, -1.1) | 68.02 | 57.66 | 54.64 | -1.2 (-1.7, -0.7)* | 64.1 |
| NH other | 28.73 | 25.52 | 17.77 | -3.2 (-4.2, -2.3) | 48.04 | 39.58 | 29.61 | -3.0 (-3.8, -2.3)* | 64.8 |
| Sex |  |  |  |  |  |  |  |  |  |
| Male | 48.60 | 40.90 | 30.34 | -2.4 (-2.7, -2.1) | 68.31 | 59.54 | 47.23 | -2.0 (-2.3, -1.7)* | 67.6 |
| Female | 41.51 | 33.77 | 28.67 | -2.1 (-2.6, -1.7) | 66.06 | 53.81 | 47.96 | -1.6 (-2.1, -1.2)* | 62.7 |
| **Age at death, 1-4 y** |  |  |  |  |  |  |  |  |  |
| Overall | 1.43 | 1.14 | 0.95 | -2.0 (-2.7, -1.2) | 1.93 | 1.53 | 1.25 | -1.8 (-2.4, -1.1)* | 74.1 |
| Race/ethnicity |  |  |  |  |  |  |  |  |  |
| NH white | 1.14 | 0.96 | 0.68 | -2.4 (-3.6, -1.2) | 1.62 | 1.29 | 0.93 | -2.3 (-3.2, -1.3)* | 73.0 |
| NH black | 2.43 | 1.76 | 1.67 | -1.8 (-3.1, -0.4) | 3.06 | 2.40 | 2.12 | -1.4 (-2.5, -0.2)* | 75.5 |
| Hispanic | 1.54 | 1.16 | 0.96 | -2.1 (-3.4, -0.8) | 1.96 | 1.53 | 1.30 | -1.7 (-2.9, -0.5)* | 74.4 |
| NH other | 1.55 | 1.26 | 1.17 | -1.2 (-3.4, 1.1) | 1.93 | 1.68 | 1.35 | -1.3 (-3.4, 0.8) | 76.1 |
| Sex |  |  |  |  |  |  |  |  |  |
| Male | 1.63 | 1.21 | 1.03 | -2.4 (-3.1, -1.6) | 2.12 | 1.53 | 1.36 | -2.2 (-2.9, -1.5)* | 75.1 |
| Female | 1.23 | 1.07 | 0.87 | -1.5 (-2.7, -0.3) | 1.73 | 1.54 | 1.14 | -1.3 (-2.4, -0.3)* | 72.8 |
| **Age at death, 5-17 y** |  |  |  |  |  |  |  |  |  |
| Overall | 0.46 | 0.3 | 0.28 | -3.0 (-3.6, -2.3) | 0.56 | 0.38 | 0.40 | -2.1 (-3.2, -0.9)* | 78.2 |
| Race/ethnicity |  |  |  |  |  |  |  |  |  |
| NH white | 0.42 | 0.30 | 0.25 | -3.3 (-4.1, -2.5) | 0.51 | 0.35 | 0.36 | -2.2 (-4.0, -0.3)* | 77.5 |
| NH black | 0.69 | 0.42 | 0.49 | -2.1 (-3.3, -1.0) | 0.78 | 0.57 | 0.64 | -1.7 (-2.7, -0.7)* | 82.8 |
| Hispanic | 0.41 | 0.20 | 0.22 | -2.6 (-4.1, -1.1) | 0.49 | 0.31 | 0.32 | -2.0 (-3.5, -0.5)* | 75.7 |
| NH other | 0.38 | 0.23 | 0.38 | -1.4 (-3.6, 1.0) | 0.61 | 0.37 | 0.46 | -1.5 (-3.5, 0.5) | 74.8 |
| Sex |  |  |  |  |  |  |  |  |  |
| Male | 0.58 | 0.33 | 0.35 | -2.9 (-3.6, -2.1) | 0.67 | 0.41 | 0.48 | -1.8 (-3.3, -0.3)* | 80.1 |
| Female | 0.35 | 0.26 | 0.21 | -3.1 (-3.8, -2.3) | 0.44 | 0.35 | 0.32 | -2.5 (-3.3, -1.7)* | 75.6 |
| **Age at death, 18-34 y** |  |  |  |  |  |  |  |  |  |
| Overall | 0.61 | 0.49 | 0.36 | -3.3 (-3.8, -2.8) | 0.77 | 0.64 | 0.53 | -2.4 (-2.9, -1.9)* | 74.4 |
| Race/ethnicity |  |  |  |  |  |  |  |  |  |
| NH white | 0.65 | 0.48 | 0.35 | -3.8 (-4.4, -3.2) | 0.80 | 0.65 | 0.56 | -2.6 (-3.2, -2.0)* | 73.3 |
| NH black | 0.71 | 0.74 | 0.50 | -2.8 (-3.8, -1.8) | 0.93 | 0.94 | 0.62 | -2.3 (-3.2, -1.3)* | 77.3 |
| Hispanic | 0.44 | 0.37 | 0.37 | -1.8 (-3.5, 0.0) | 0.59 | 0.47 | 0.45 | -1.5 (-2.9, 0.0) | 76.1 |
| NH other | 0.44 | 0.40 | 0.18 | -1.6 (-3.7, 0.6) | 0.54 | 0.46 | 0.33 | -1.0 (-2.7, 0.7) | 73.2 |
| Sex |  |  |  |  |  |  |  |  |  |
| Male | 0.67 | 0.66 | 0.49 | -2.6 (-3.3, -1.9) | 0.88 | 0.88 | 0.71 | -1.8 (-2.4, -1.2)* | 74.4 |
| Female | 0.56 | 0.31 | 0.23 | -4.7 (-5.4, -4.0) | 0.67 | 0.40 | 0.34 | -3.6 (-4.3, -3.0)* | 74.3 |
| **Age at death, 35-49 y** |  |  |  |  |  |  |  |  |  |
| Overall | 0.64 | 0.48 | 0.38 | -2.2 (-2.7, -1.7) | 0.91 | 0.76 | 0.61 | -1.6 (-2.1, -1.2)* | 65.2 |
| Race/ethnicity |  |  |  |  |  |  |  |  |  |
| NH white | 0.69 | 0.54 | 0.46 | -1.5 (-2.1, -0.8) | 0.95 | 0.83 | 0.73 | -1.0 (-1.5, -0.5)* | 64.8 |
| NH black | 0.70 | 0.45 | 0.36 | -2.5 (-3.7, -1.2) | 1.02 | 0.78 | 0.51 | -3.4 (-8.3, 1.6) | 68.2 |
| Hispanic | 0.39 | 0.35 | 0.28 | -2.7 (-4.1, -1.2) | 0.64 | 0.56 | 0.46 | -1.7 (-2.9, -0.4)* | 63.5 |
| NH other | 0.32 | 0.25 | 0.12 | -2.4 (-4.6, -0.2) | 0.48 | 0.33 | 0.21 | -1.4 (-3.1, 0.4) | 66.5 |
| Sex |  |  |  |  |  |  |  |  |  |
| Male | 0.69 | 0.54 | 0.46 | -1.7 (-2.3, -1.1) | 1.02 | 0.89 | 0.77 | -1.1 (-1.5, -0.6)* | 63.6 |
| Female | 0.60 | 0.42 | 0.32 | -2.8 (-3.6, -2.0) | 0.81 | 0.63 | 0.44 | -2.4 (-3.2, -1.6)* | 67.2 |
| **Age at death, 50-64 y** |  |  |  |  |  |  |  |  |  |
| Overall | 0.65 | 0.46 | 0.4 | -2.4 (-3.4, -1.4) | 1.04 | 0.78 | 0.80 | -1.0 (-1.9, -0.1)* | 57.1 |
| Race/ethnicity |  |  |  |  |  |  |  |  |  |
| NH white | 0.67 | 0.47 | 0.45 | -1.9 (-3.1, -0.8) | 1.08 | 0.81 | 0.88 | -1.1 (-1.9, -0.2)* | 57.3 |
| NH black | 0.74 | 0.70 | 0.37 | -3.5 (-5.6, -1.3) | 1.26 | 1.07 | 0.84 | -2.4 (-4.1, -0.6)* | 55.8 |
| Hispanic | 0.38 | 0.31 | 0.24 | -2.8 (-4.9, -0.7) | 0.58 | 0.44 | 0.49 | -1.7 (-3.2, -0.1)* | 57.8 |
| NH other | 0.47 | 0.19 | 0.20 | -3.4 (-6.3, -0.3) | 0.65 | 0.38 | 0.41 | -1.5 (-3.4, 0.5) | 57.6 |
| Sex |  |  |  |  |  |  |  |  |  |
| Male | 0.72 | 0.53 | 0.46 | -1.4 (-2.2, -0.5) | 1.19 | 0.89 | 0.98 | -0.4 (-1.4, 0.7) | 54.0 |
| Female | 0.59 | 0.41 | 0.35 | -3.0 (-3.7, -2.2) | 0.91 | 0.69 | 0.63 | -2.4 (-3.2, -1.6)* | 61.1 |
| **Age at death, ≥65 y** |  |  |  |  |  |  |  |  |  |
| Overall | 1.45 | 0.83 | 0.64 | -4.2 (-4.9, -3.5) | 2.31 | 1.48 | 1.26 | -3.3 (-4.0, -2.5)* | 55.5 |
| Race/ethnicity |  |  |  |  |  |  |  |  |  |
| NH white | 1.50 | 0.87 | 0.69 | -3.9 (-4.6, -3.2) | 2.38 | 1.55 | 1.41 | -2.8 (-3.6, -2.0)* | 55.3 |
| NH black | 1.39 | 0.86 | 0.43 | -5.0 (-6.8, -3.2) | 2.14 | 1.28 | 0.71 | -5.7 (-8.7, -2.6)* | 58.6 |
| Hispanic | 1.15 | 0.30 | 0.50 | -5.4 (-9.1, -1.6) | 1.57 | 0.91 | 0.76 | -4.7 (-6.5, -2.9)* | 55.3 |
| NH other | 0.42 | 0.65 | 0.52 | -4.1 (-6.4, -1.7) | 1.49 | 1.16 | 0.78 | -4.9 (-6.8, -2.9)* | 56.4 |
| Sex |  |  |  |  |  |  |  |  |  |
| Male | 1.27 | 0.81 | 0.63 | -3.7 (-5.2, -2.2) | 2.11 | 1.49 | 1.35 | -2.5 (-3.7, -1.2)* | 52.6 |
| Female | 1.59 | 0.84 | 0.65 | -4.5 (-5.7, -3.2) | 2.44 | 1.47 | 1.19 | -3.8 (-4.9, -2.7)* | 57.5 |

Footnote: AAPC, average annual percent change; CI, confidence interval
Mortality rates are calculated as the number of deaths per 100,000 population for all age groups one year or older and as the number of deaths per 100,000 live births for the infant (<1 year) age group.
Mortality rates for "all ages" are age-standardized; mortality rates for individual age groups are crude (age-specific).

**Supplemental Figure 1.**
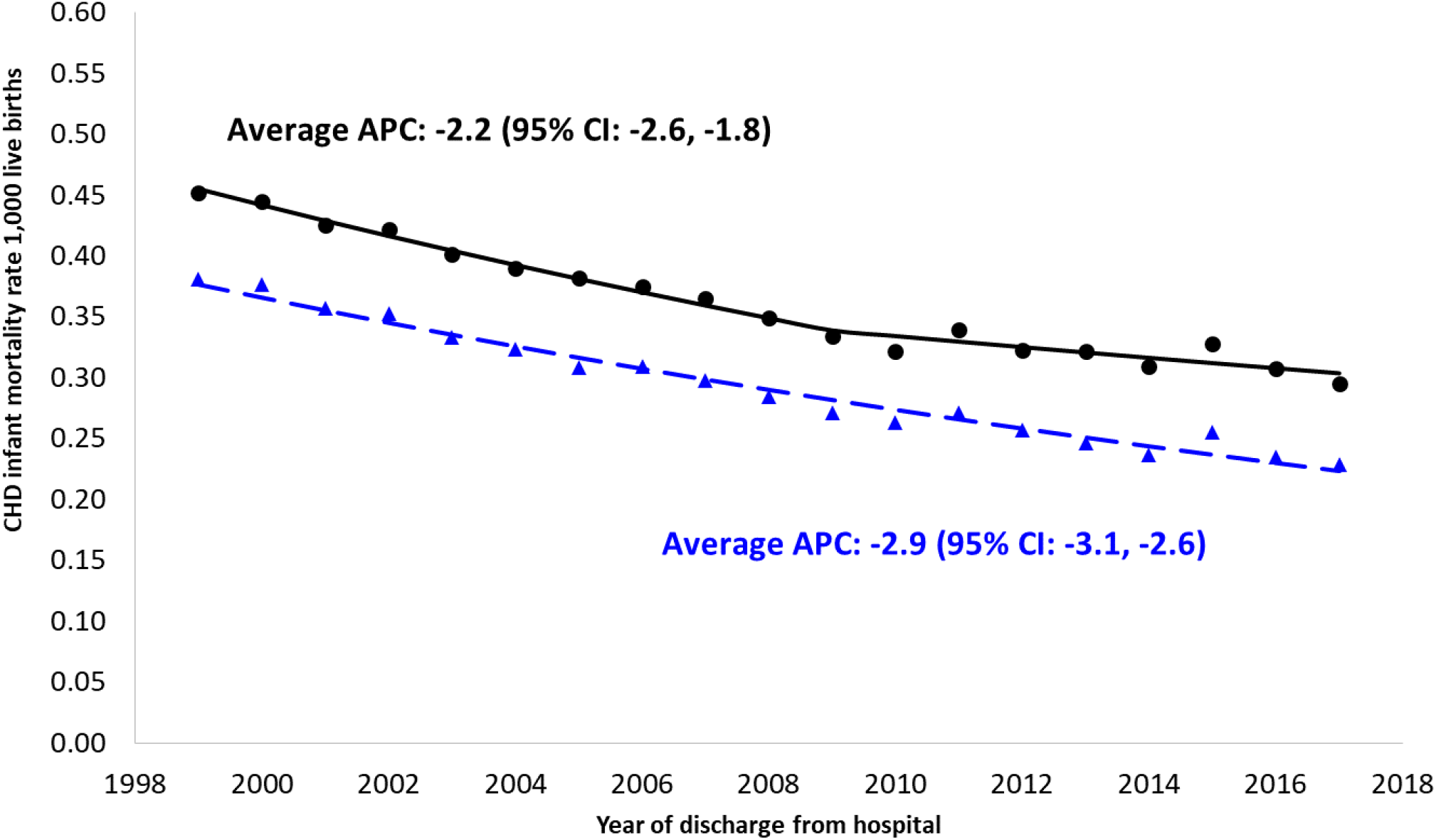
Temporal trends in infant mortality due to CHD, with and without the exclusion of deaths with prematurity, extracardiac defects, and genetic syndromes as a contributing case, 1999-2017. Footnotes: APC, annual percent change The solid black line includes all deaths with a CHD listed as the underlying cause of death. The blue dashed line excludes those cases with prematurity, extracardiac defects, or genetic syndromes listed as a contributing cause of death.

## Notes

### Competing Interest Statement

The authors have declared no competing interest.

